# Functional and molecular characterization of suicidality factors using phenotypic and genome-wide data

**DOI:** 10.1101/2022.10.26.22281550

**Authors:** Andrea Quintero Reis, Brendan A Newton, Ronald Kessler, Renato Polimanti, Frank R Wendt

## Abstract

Genome-wide association studies (GWAS) of suicidal thoughts and behaviors support the existence of genetic contributions. Continuous measures of psychiatric disorder symptom severity can sometimes model polygenic risk better than binarized definitions. We compared two severity measures of suicidal thoughts and behaviors at the molecular and functional levels using genome-wide data. We used summary association data from GWAS of four traits analyzed in 122,935 individuals of European ancestry: *thought life was not worth living* (TLNWL), *thoughts of self-harm, actual self-harm*, and *attempted suicide*. The fifth trait, suicidality, was constructed with phenotypically as an aggregate of these four traits and genetically using Genomic Structural Equation modeling. Suicidality and S-factor were compared at the level of SNP-heritability (*h*^*2*^), genetic correlation, partitioned *h*^*2*^, effect size distribution, transcriptomic effects in the brain, and cross-population polygenic scoring (PGS). The S-factor had good model fit (*χ*^2^=0.21, AIC=16.21, CFI=1.00, SRMR=0.024). Suicidality (*h*^*2*^=7.6%) had higher *h*^*2*^ than the S-factor (*h*^*2*^=2.54, P_diff_ =4.78×10^-13^). Although the S-factor had a larger number of non-null susceptibility loci (π_c_=0.010), these loci had small effect sizes compared to those influencing suicidality (π_c_ =0.005, P_diff_ =0.045). The *h*^*2*^ of both traits was enrichment for conserved biological pathways. The *r*g and *ρ*_GE_ support highly overlapping genetic and transcriptomic features between suicidality and the S-factor. PGS using European-ancestry SNP effect sizes strongly associated with TLNWL in Admixed Americans: Nagelkerke’s *R*^*2*^=8.56%, P=0.009 (PGS_suicidality_) and Nagelkerke’s *R*^*2*^=7.48%, P=0.045 (PGS_S-factor_). An aggregate suicidality phenotype was statistically more heritable than the S-factor across all analyses and may be more informative for future study genetic designs than individual suicidality indicator traits.

## INTRODUCTION

Death by suicide is responsible for more than 700,000 deaths per year.^1^ The World Health Organization invested in global advocacy and awareness programs towards reducing stigma and increasing access to care. However, death by suicide ranks as the fourth leading cause of death among teens and young adults.^2^ Twin, family, and adoption studies show a heritability (i.e., phenotypic variation explained by genetic differences) of suicidal thoughts and behaviors between 30-50%.^3^

Large genome-wide association studies (GWAS) of individual thoughts and behaviors associated with death by suicide demonstrate small but robust heritability estimates attributed to common genetic variation (*h*^*2*^): 1.25% for attempted suicide (AS) in the Million Veteran Program (MVP),^4^ 1.9%-4.6% in a Danish study of AS with and without considering mental health diagnoses,^5^ 5.7%-6.8% in two large meta-analyses of AS lead by the International Suicide Genetics Consortium.^1, 6^ Among individuals with co-occurring diagnoses of major depressive disorder, bipolar disorder, and/or schizophrenia, the Psychiatric Genomics Consortium reported null *h*^*2*^ for AS.^7^ Taken together, these studies support a common genetic component to suicidal thoughts and behaviors that may be independent of other mental health diagnoses.

Other suicidal behaviors occur well before AS and may include ideation and planning the attempt to end one’s life. Though death by suicide is not a diagnosis, these thoughts and behaviors may be modeled as a severity continuum termed ‘suicidality’ analogous to symptom severity measures used for formal DSM diagnoses. Among GWAS of psychiatric disorders and conditions associated with AS and death by suicide, continuous measures of symptom severity appear to better model polygenic risk than binarized case-control items, leading to increased statistical power in gene discovery.^8, 9^ This is especially true for traits strongly correlated with suicidal thoughts and behaviors such as posttraumatic stress disorder ^8^ and major depressive disorder (MDD).^10^ It therefore stands to reason that a suicidality measure capturing continuous variation across suicidal thoughts and behaviors would be more statistically powerful than any individual dichotomized definition. Strawbridge, *et al*. reported a genome-wide association study of one definition of suicidality in the UK Biobank and reported an *h*^*2*^ of 7.6% in a sample substantially smaller than the most contemporary meta-analyses of SA.^1, 4, 6, 7, 11^

Though showing higher *h*^*2*^ than individual thoughts and behaviors associated with death by suicide, aggregating these items at the phenotype level may induce phenotypic heterogeneity that limits the potential discovery of genome-wide significant loci and biological processes relevant for discrete, yet related, behaviors. A primary limitation of some recently introduced phenotypically aggregated suicidality measures is the equal contribution of each questionnaire item to the final suicidality rating regardless of the heritability of each item or the relationship between items. Genomic structural equation modeling (gSEM^12, 13^) is a multivariate method that permits building factor structure(s) that account for the heritability of each indicator and the relationship between indicators. For example, gSEM has been used previously to describe how the 10-item Alcohol Use Disorder Identification Test reflects a correlated two-factor structure of problematic use and consumption.^14^

This study asked whether there is any benefit to studying a gSEM derived “S-factor” in addition to a questionnaire-derived suicidality measure to inform suicide biology. We report *h*^*2*^ differences between suicidality and S-factor and compare these trait definitions on the basis of genetic correlation with other psychopathologies and mental health diagnoses, functional enrichment underlying their *h*^*2*^ estimates, and transcriptomic signatures across various relevant brain regions. Our findings reinforce the greater statistical power of a phenotypically-derived suicidality factor and demonstrate a systematic reduction of signal in all analyses when analyzing the S-factor.

## METHODS

### S-factor Modeling

Using gSEM,^12, 13^ common factor GWAS was performed on the S-factor linking four traits describing the thoughts and behaviors associated with death by suicide and range in severity from ideation to attempt to end one’s life. The four common factor indicators were questions from the UK Biobank (UKB) self-harm behaviors section of the online Mental Health Questionnaire and have been previously described by Strawbridge, *et al*.:^11^ *thought life was not worth living* (TLNWL), *thoughts of self-harm or suicide* (TSH), *actual self-harm* (ASH), and *attempted suicide* (AS). Note that participants could respond to ASH with “yes” for deliberate acts of self-harm whether or not they intended to end their own lives.

Multivariable linkage disequilibrium score regression (LDSC) was used to obtain a genetic covariance and corresponding sampling matrix based on a European ancestry linkage disequilibrium reference panel reflecting the 1000 Genomes Project EUR superpopulation. All factor modeling used diagonally weighted least squares estimation and promax rotation.

### Trait Description

The UKB is a population-based cohort of >500,000 participants with deep phenotyping of lifestyle factors, mental and physical health outcomes, anthropometric measurements, and other traits. Our analysis used the self-harm behavior GWAS summary data from unrelated European ancestry participants adjusted for age, sex, genotyping chip, and within ancestry genetic principal components. TLNWL (UKB Field ID 20479) and TSH (UKB Field ID 20485) asked participants to respond “No,” “Yes, once,” or “Yes, more than once” to questions about thought/contemplation of self-harm. These items were dichotomized into “no” and “yes” for GWAS.^11^ ASH (UKB Field ID 20480) and AS (UKB Field ID 20483) asked participants to respond with “no” or “yes” to questions about actual self-harm behavior. These four items also were aggregated into a single ordinal trait termed “suicidality” such that participants responding “no” to all four questions were assigned “0” and each “yes” increased the participants’ suicidality score up to 4 (most severe). UKB participants with death-by-suicide ICD codes X60-X84 (classified as intentional self-harm) were excluded from GWAS. Further description of these variables has been published previously.^11, 15, 16^

### Linkage Disequilibrium Score Regression (LDSC)

LDSC was used to estimate the *h*^*2*^-SNP of the *S*-factor based on the 1000 Genomes Project European ancestry reference panel. Stratified-LDSC (S-LDSC) was implemented in GenomicSEM for >51 genomic annotations (baseline annotation v2.2 with flanking and continuous annotations excluded) related to allele frequency strata, genomic conservation, evolutionary selective pressure, epigenomic regulatory sites, etc.^17-20^ The major histocompatibility complex region was excluded from these analyses due to its complex linkage disequilibrium structure.

LDSC also was used to estimate the genetic correlation (*r*_*g*_) between suicidality and the *S*-factor relative to various suicide-associated traits and risk factors including large GWAS of psychiatric disorders. These were: TLNWL, TSH, ASH, and AS reported by Strawbridge, et al. ^11^; suicide attempt among bipolar disorder, schizophrenia, and major depression cases from Mullins, et al. ^7^; psychiatric disorder GWAS from the Psychiatric Genomics Consortium including ADHD ^21^, anorexia nervosa ^22^, obsessive compulsive disorder ^23^, schizophrenia ^24^, Tourette syndrome ^25^ and the Million Veteran Program including problematic alcohol use ^26^, posttraumatic stress disorder and its symptom domains ^8^, broad depression ^10^, and generalized anxiety disorder ^27^; personality domains from the Genetics of Personality Consortium including extraversion, agreeableness, conscientiousness, openness ^28^; and other related variables from the Social Science Genetic Association Consortium including subjective well-being ^29^, neuroticism ^29^, risky behavior ^30^, risk tolerance ^30^, cognitive performance ^31^, education years ^31^, and educational attainment ^31^.

### Effect size distribution

The R package GENESIS^32^ was used to estimate common variant effect size distributions for suicidality and the S-factor. Effect size distributions are characterized by three statistics: π_c_ describes the proportion of susceptibility SNPs, σ^2^ describes the variance in effect size for non-null SNPs, and α describes residual effects not captured by the variance of effect-sizes such as population stratification, underestimated effects of extremely small effect size SNPs, and/or genomic deflation. We performed 2-component modeling to specify the effect of non-null SNPs.^20, 32, 33^ GWAS data were filtered to (i) include only HapMap3 SNPs (excluding the major histocompatibility complex due to its complex linkage disequilibrium structure), (2) exclude SNPs with Z^2^>80, and (3) exclude SNPs with effective samples sizes less than 0.67-times the 90th percentile of the total sample.

We also included GWAS data for height^34^ and broad depression.^10^ Height was used as a model trait with broad effect size distribution representing a relatively large proportion of non-null SNPs with relatively large effect sizes. MDD was used as a model trait with narrow effect size distribution representing a relatively small proportion of non-null SNPs with relatively small effect sizes.

### GTEx v8 Tissue Enrichment

Tissue transcriptomic profile enrichment was evaluated using Multi-marker Analysis of GenoMic Annotation (MAGMA).^35^ To identify tissue effects of each phenotype, gene-property analyses were applied with Functional Mapping and Annotation (FUMA) to test relationships between tissue-specific gene expression profiles and disease-gene associations.^36^

### Transcriptome-wide Association Studies

Summary-based transcriptome-wide association studies (TWAS) of suicidality and the S-factor were performed using the GTEx v8 TWAS expression weights for cerebellar hemisphere, cerebellum, hippocampus, and hypothalamus. Gene expression weights for 6,091 features of the cerebellar hemisphere are estimated in 157 individuals, for 7,272 features of the cerebellum are estimated in 188 individuals, for 3,547 features of the hippocampus is estimated in 150 individuals, and for 3,543 features of the hypothalamus are estimated in 156 individuals. FUSION^37^ was used to perform TWAS of suicidality and all four S-factor indicators. TWAS of the S-factor was performed using gSEM and the TWAS summary association data from FUSION for each S-factor indicator. Multiple testing correction was applied using a Bonferroni threshold per tissue (P<8.21×10^-6^=0.05/6,091 genes for cerebellar hemisphere; P<6.88×10^-6^=0.05/7,272 genes for cerebellum; P<1.41×10^-5^=0.05/3,547 genes for hippocampus; P<1.41×10^- 5^=0.05/3,543 genes for hypothalamus). RHOGE was used to estimate the genome-wide genetic correlation between suicidality and the S-factor as a function of predicted *cis* gene expression effects on each trait.^38^

### Cross-ancestry Translation of European-ancestry Polygenic Risk

We applied cross-ancestry polygenic scoring in the UK Biobank (Application Number 58146) to evaluate how findings from individuals of European ancestry extend to diverse communities. We derived suicidality in five additional groups as described previously: African (N=876), Admixed American (N=256), Central/South Asian (N=1,106), East Asian (N=599), and Middle Eastern (N=269).^11, 16^ Ancestry groups were defined using a random forest classifier based on genetic principal components relative to a combined reference panel from the 1000 Genomes Project Phase III and the Human Genome Diversity Project. This procedure is described in detail at the Pan-Ancestry UK Biobank web-page: https://pan.ukbb.broadinstitute.org/docs/qc.

Suicidality and S-factor polygenic scores (PGS) with continuous shrinkage were calculated for individuals from each ancestry group using PRS-CS.^39^ PRS-CS is a Bayesian polygenic prediction method that imposes continuous shrinkage priors on SNP effect sizes. LD-independent SNPs were selected based on the UK Biobank European ancestry reference panel. We further required that each SNP have a minor allele frequency greater than 5% in the target ancestry group. Generalized linear models associating suicidality with suicidality and S-factor polygenic scores included age, age^2^, sex×age, sex×age^2^, and ten within-ancestry genetic principal components. We also tested the association of polygenic scores with each suicidality indicator trait (note that AS had too few observations to test) and depression (endorsement of either UKB Field ID 2090 or 2100),^40^ total neuroticism score (inverse rank normalized, UKB Field ID 20127), and standing height (in centimeters; UKB Field ID 50).

## RESULTS

### The S-factor Structure

Using GenomicSEM, we constructed a common factor model that we refer to as the *S*-factor as it reflects the multivariate effects of the thoughts and behaviors proceeding death by suicide. We considered four indicators in the *S*-factor: *thought life not worth living* = “TLNWL,” *thoughts of self-harm* = “TSH,” *actual self-harm* = “ASH,” and *attempted suicide* = “AS.” TLNWL, TSH, and ASH had significant non-zero *h*^*2*^ (Table S1) but AS did not (*h*^*2*^=3.34%, P=0.099) so we considered two *S*-factor models: model-1 included TLNWL, TSH, ASH, and AS (Figure 1A) and model-2 included only TLNWL, TSH, and ASH. Genetic correlations (*r* g s) for each pair of indicators are shown in Table S2. Model-1 (*χ*^2^(2)=0.21, AIC=16.21, CFI=1.00, SRMR=0.024) had superior fit statistics relative to model-2 (*χ*^2^(2)=3.14, AIC=19.14, CFI=0.999, SRMR=0.025) and was chosen for all subsequent *S*-factor analyses (Table S3). TSH was the indicator most strongly loaded onto the *S*-factor (standardized loading = 1 ± 0.17).

**Fig 1.**
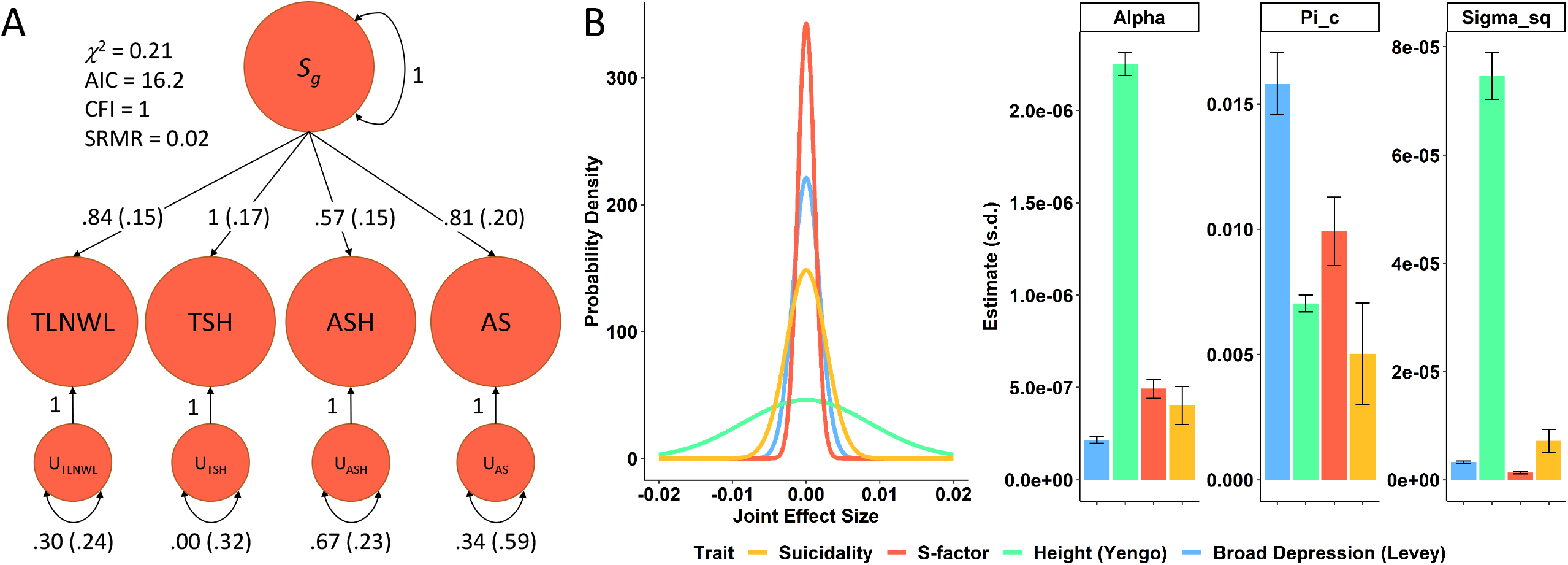
Factor structure and genetic architecture of the S-factor. (A) The four thoughts and behaviors associated with death by suicide (*thought life not worth living* = TLNWL, *thoughts of self-harm* = TSH, *actual self-harm* = ASH, and *attempted suicide* = AS) fit a single common factor (*S*-factor fit statistics are shown in the top left corner). The TSH indicator loading was constrained to 1; Table S3 shows indicator loadings before and after Heywood-case correction. (B) Genetic effect size distribution and associated statistics of the S-factor relative to suicidality using height and broad depression as comparative traits with relatively large and small proportions of relatively high effect size SNPs, respectively.

### Genetic Architecture of the S-factor

There were no loci associated with the *S*-factor at the level of genome-wide significance (GWS, P<5×10^-8^; Figure 1). All three suicidality loci from Strawbridge, *et al*.^11^ were nominally replicated in the *S*-factor GWAS (Table S4) with no significant difference in effect size between the two studies.

We quantified several metrics of genome-wide polygenicity (Figure 1B) using GENESIS.^41^ Relative to suicidality (π_c_=0.005±0.002), the S-factor has a significantly higher proportion of non-null susceptibility SNPs (π_c_=0.010±0.001, P_diff_=0.045). However, the suicidality effect size distribution is broader than that of the S-factor, suggesting that suicidality SNPs effect sizes are generally greater and may require smaller samples sizes to detect by GWAS.^11^ When projected sample sizes reach 500,000 the S-factor GWAS is estimated to remain uninformative while the suicidality GWAS should yield 60 GWS SNPs (95% CI: 15-139). At projects of 1-million individuals, the S-factor remains relatively uninformative (4 GWS SNPs, 95% CI: 0-18). Only when projected to 5-million individuals does the S-factor GWAS become an informative source of associated loci (539 GWS SNPs, 95% CI: 208-1117); however, the suicidality GWAS remained the more lucrative study in terms of susceptibility loci discovered at all projected sample sizes (Table S5).

### Heritability Comparisons

The *h*^*2*^-SNP of the S-factor was 2.54% (P=1.72×10^-12^) which is significantly lower than the *h*^*2*^-SNP for a pooled suicidality phenotype (*h*^*2*^-SNP=7.6%, P_diff_ =4.78×78^-13^).^11^ Though different with respect to phenotypic variance explained by common genetic variation, the *r*_*g*_ between the S-factor and suicidality is almost perfect (*r* g =0.996, P<9.21×21^-308^).

S-LDSC was applied to quantify the overlap between genomic annotation contributions to *h*^*2*^-SNP in the suicidality and *S*-factor GWAS. We partitioned the *h*^*2*^-SNP of the S-factor three ways: with LDSC, with gSEM S-LDSC using all indicators, and with gSEM S-LDSC removing the least well-powered indicator (AS). This approach permitted comparison across methods for robust detection of enriched genomic categories using the various strengths of each method (e.g., gSEM S-LDSC reports a Z-smooth value quantifying the degree of smoothing applied to the data with clear guidelines for enrichment interpretation given these values). The most consistently enriched annotation described sites conserved across primates as measured by PhastCons 46-way alignment (*converved_primate_phastcons46way*, mean enrichment=19.80±5.68, Figure 2 and Table S6). Regardless of trait definition or partitioning method applied, there were no significant differences in genomic enrichment (P≥0.147).

**Fig 2.**
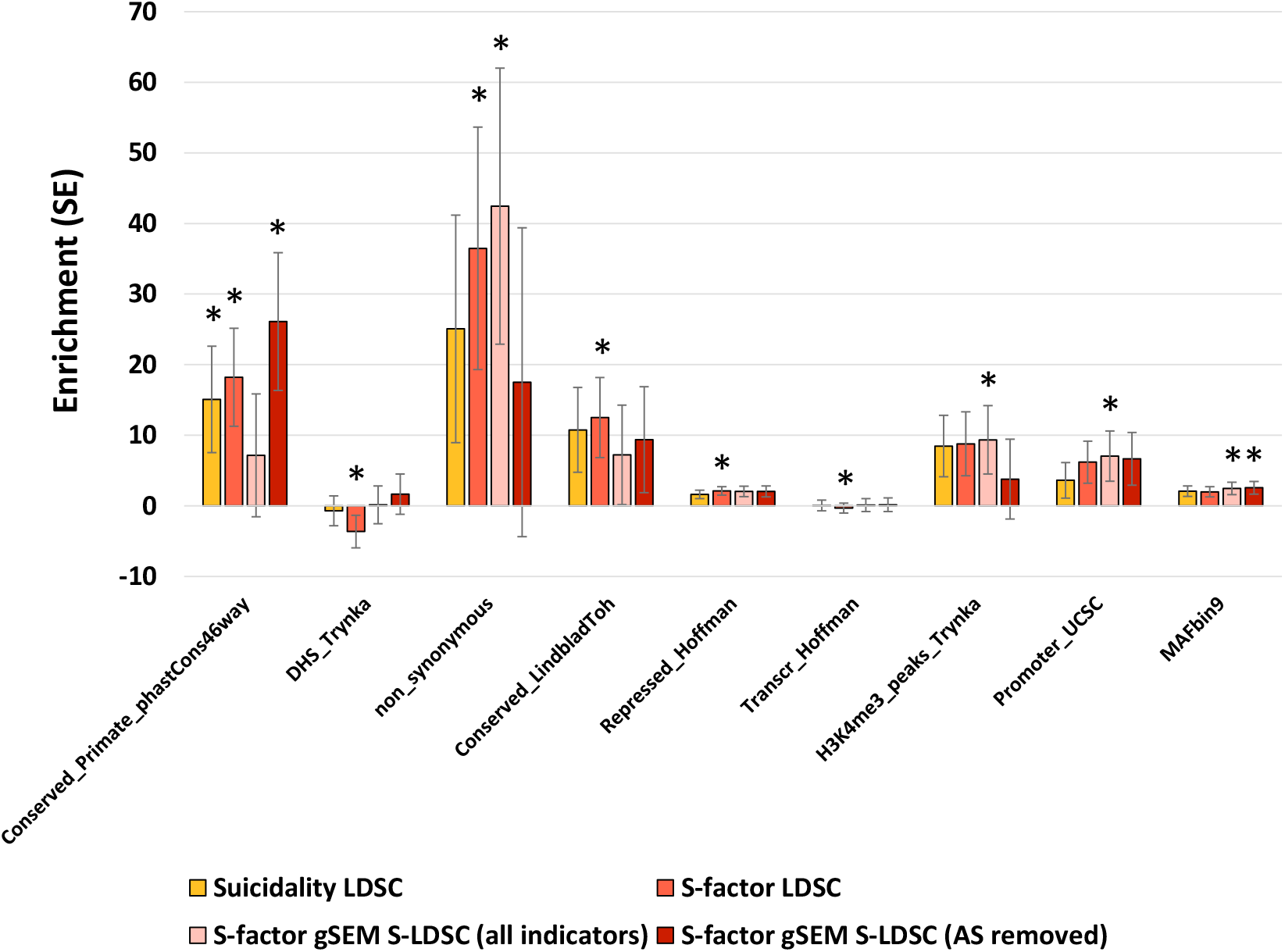
Genomic annotation enrichment. Enrichment of 9 genomic annotations at least nominally enriched (asterisks indicate P<0.05) in the GWAS of suicidality and/or the S-factor. Genomic annotations have been described previously.^17, 18, 20^ All genomic annotation enrichments are provided in Table S6.

### Genetic Correlation

We next compared *r*_*g*_ estimates of suicidality and the S-factor relative to 32 mental health traits, including other genetic assessments of the thoughts and behaviors associated with death by suicide. Because of the high genetic overlap between the S-factor and suicidality, the *r*_*g*_ estimates with other mental health traits were nearly identical (Figure 3, adjusted *R*^*2*^=0.984, P=8.55×55^-188^). Though not significantly different, the largest magnitude of difference in *r*_*g*_ estimates stems from comparisons with TLNWL (*r*_*g*_ with suicidality=0.870, *r*_*g*_ with S-factor=0.941, P_diff_=0.284).

**Fig 3.**
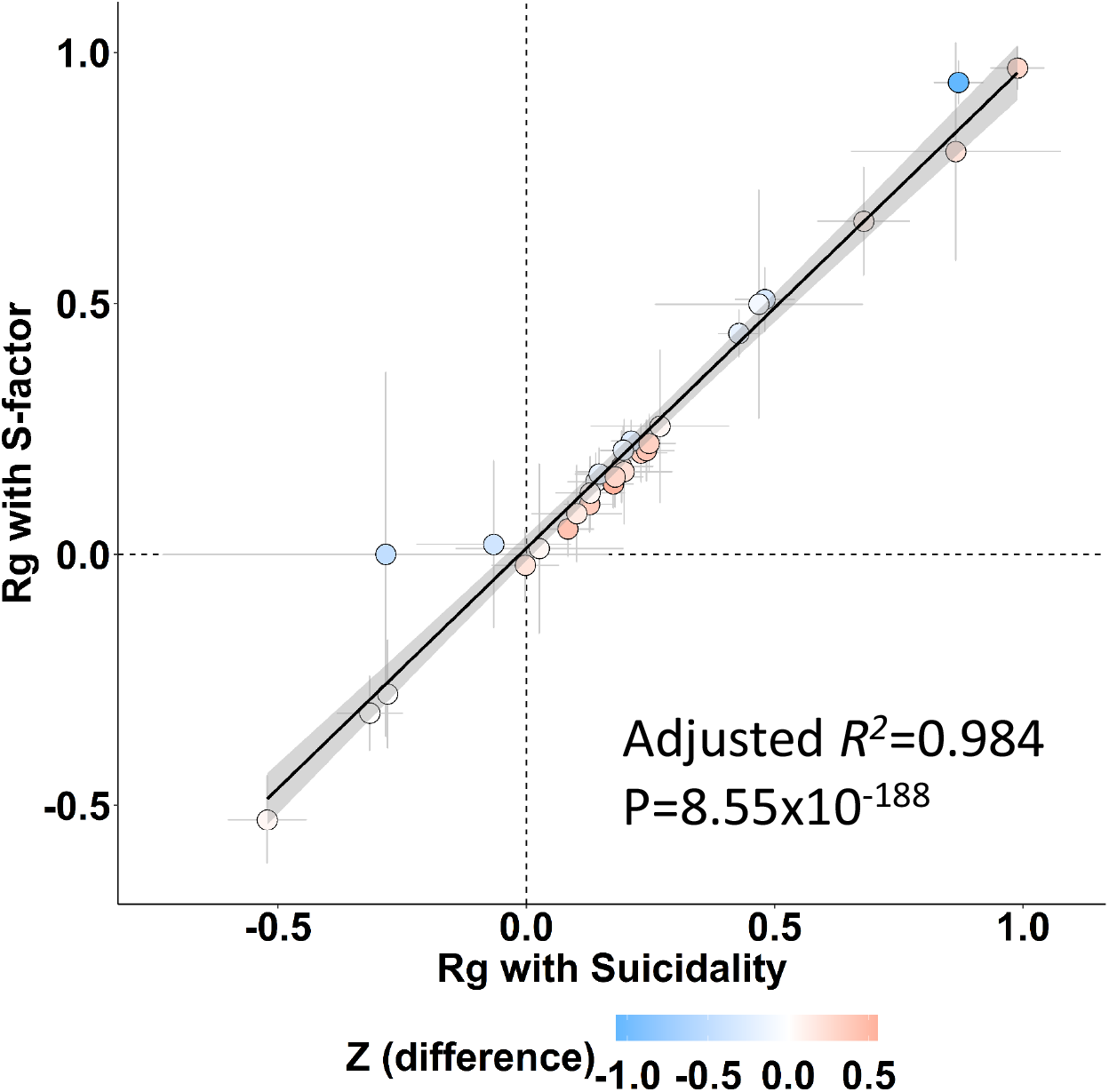
Genetic correlation estimates for suicidality and S-factor and mental health/psychopathology traits. Each data point reflects a single mental health/psychopathology trait genetically correlated with suicidality and the S-factor (Table S7). Error bars denote the standard error associated with each point estimate. The adjusted-*R*^*2*^ reflects a linear prediction of S-factor *r*_g_ with no intercept and weighted by the standard error of each *r*_*g*_ with suicidality.

### Comparison of Brain Region Transcriptomic Effects

Consistent with prior findings from this study, tissue transcriptomic profile enrichments for suicidality and the S-factor are highly correlated (R^2^=0.861, P=3.265×265^-34^). Though the S-factor GWAS yielded two significant tissue transcriptomic profile enrichments, there was no difference in effect size for the enrichments derived from suicidality or the S-factor. Gene expression weights from four brain tissues were used for TWAS comparisons of suicidality and the S-factor due to their significant enrichments in the suicidality GWAS: cerebellar hemisphere, cerebellum, hippocampus, and hypothalamus (Table S8).

Though no gene reached GWS in any of the TWAS performed (Table S9), the genetic correlation between genetically predicted gene expression effects underlying suicidality and the S-factor in each tissue was high: *ρ*_GE_ =0.990, P=1.31×31^-296^ considering cerebellar hemisphere weights; *ρ*_GE_ =0.991, P<9.21×21^-308^ considering cerebellum weights; *ρ*_GE_ = =0.991, P=8.96×96^-175^ considering hippocampus weights; *ρ*_GE_ =0.992, P=1.81×81^-193^ considering hypothalamus weights. The most significant protein-coding gene expression effects discovered from the more powerful suicidality TWAS were *RBM26* in the cerebellar hemisphere (suicidality Z=3.95, P=7.53×53^-5^; S-factor Z=2.78, P=0.005) and *COLQ* in the hippocampus (suicidality Z=-3.90, P=9.53×53^-5^; S-factor Z=3.12, P=0.001).

### Cross-population Polygenic Scoring

Using SNP effect sizes estimated from large European-ancestry GWAS, PGS for suicidality and the S-factor in diverse ancestries were highly correlated (minimum *Pearson’s r*=0.726, P=3.25×25^-214^ in AFR; maximum *Pearson’s r*=0.824, P=1.92×92^-121^ in MID; Figure 4a). PGS for suicidality associated with suicidality (*R*^*2*^=11.1%, P=0.017), TLWNL (Nagelkerke’s *R*^*2*^=8.56%, P=0.009), and ASH (Nagelkerke’s *R*^*2*^=14.3%, P=0.034) in the AMR population; PGS for the S-factor associated with TLNWL in AMR (Nagelkerke’s *R*^*2*^=7.48%, P=0.045) and with ASH in MID (Nagelkerke’s *R*^*2*^=7.71%, P=0.046; Table S10 and Figure 4b). PGS for the S-factor also associated with neuroticism scores and depression in several diverse populations but the analogous test with suicidality PGS were generally not significant. However, there were no differences in effect size for the PGS regardless of the GWAS used to train them. In the CSA population, suicidality (Nagelkerke’s *R*^*2*^=4.39%, P=0.048) and S-factor (Nagelkerke’s *R*^*2*^=4.31%, P=0.048) PGS both associated with depression (Table S10). As a null control, we observed no relationship between PGS for suicidality or S-factor and height.

**Figure 4.**
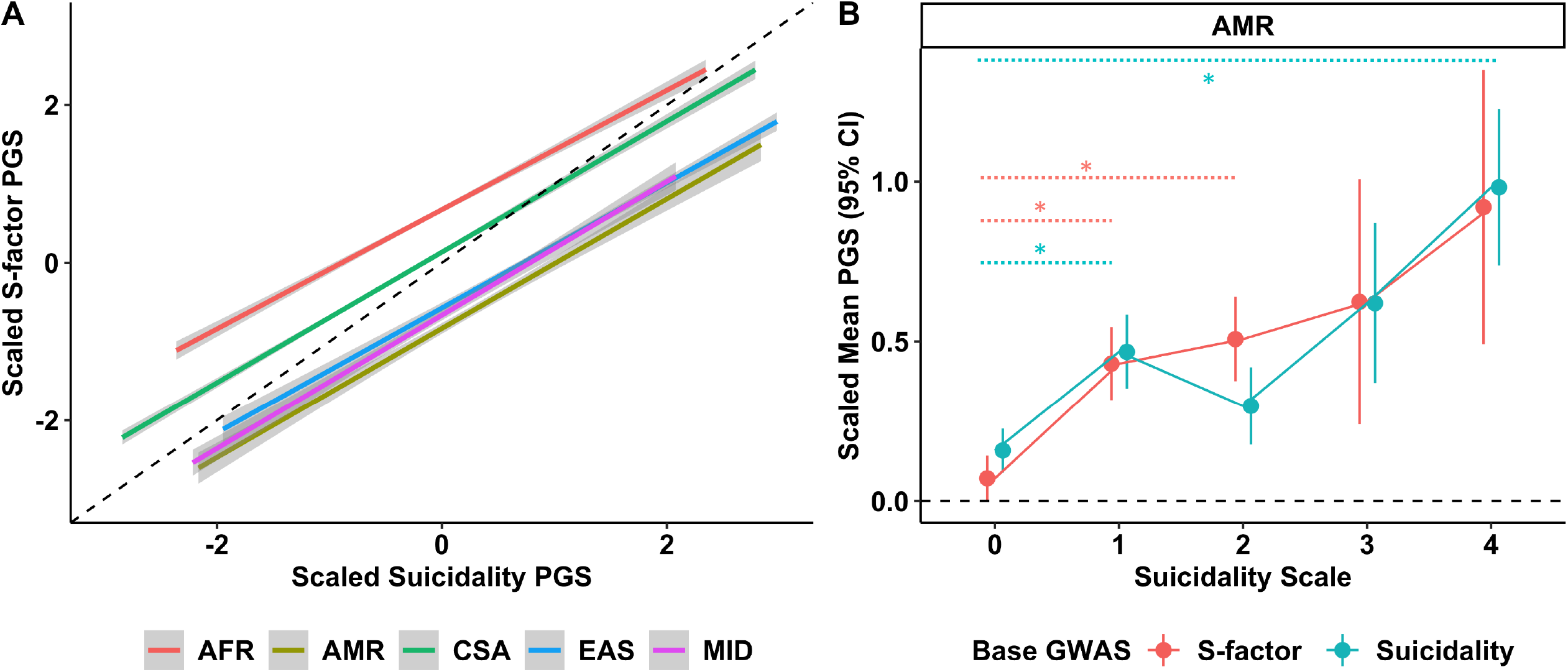
Cross-population polygenic scoring. (A) Linear relationships between suicidality and S-factor polygenic scores estimated using European ancestry GWAS summary association data in five diverse ancestry groups. (B) Portability of suicidality and S-factor polygenic scores into Admixed American suicidality data from the UK Biobank. Asterisks indicate a significant difference in polygenic score relative to the control (suicidality scale = 0).

## DISCUSSION

The use of continuous or ordinal phenotypes for GWAS of psychiatric disorders and related psychopathologies has proven a powerful way to identify risk loci and related biological pathways underlying common diagnoses.^8-10^ This approach has been previously applied to studies of suicidality that apply an equal weight to each questionnaire item.^11^ In other words, regardless of the relationship between questionnaire items or the sensitivity of those items for an outcome, each contributed equally to the outcome. We compared this approach to capturing a suicidality phenotype to gSEM-derived S-factor which explicitly considers the relationship between each indicator variable in the model.

All four suicidal thoughts and behaviors significantly loaded onto the observed S-factor. Relative to epidemiological data supporting attempted suicide as the leading predictor of future death by suicide, the genetic component of the S-factor was most associated with TSH. AS was the least heritable indicator in this study but removing AS from the S-factor reduced model fit suggesting that this indicator is relevant on the genetic level. Therefore, AS may require larger sample sizes to become highly relevant to the S-factor structure. Instead, we suspect the major contribution of TSH to the S-factor stem from a balance between (i) heritability and power and (ii) increased specificity for suicidal thoughts and behaviors. TLNWL was the most powerful indicator GWAS but has previously shown extremely high phenotypic and genetic correlation with MDD (*r* g =0.46) and neuroticism (*r* g =0.56)^11^ and is even a component of the Personal Health Questionnaire 9-item measure of depressive symptoms.^42^

The potential for risk locus discovery in GWAS of suicidality and the S-factor produced the most notable differences. Though the S-factor had a significantly larger number of non-null SNPs, these loci required substantially larger sample sizes to detect their relatively small effect sizes. It would require an estimated 5-million individuals with a similar prevalence of S-factor indicator endorsement to yield GWS SNPs in numbers already surpassed in GWAS of correlated traits like major depressive disorder^10^ and schizophrenia.^43^ The phenotypically aggregated suicidality item had an earlier return on investment producing hundreds of GWS SNPs with cohorts ranging from 500-thousand and 1-million participants. Furthermore, suicidality had a significantly higher *h*^*2*^ estimate suggesting this phenotype is more informative for inferring relevant biology through downstream *in silico* analyses. This is reinforced by a lack of significant differences in SNP effect sizes between the GWAS of suicidality and the S-factor and may stem from utilizing a linear regression for the suicidality GWAS relative to the individual logistic regressions performed for each of the UKB items contributing to the *S*-factor GWAS. The application of linear models to an ordinal trait like suicidality complicates the interpretation of how GWS loci increase or decrease risk for such thoughts and behaviors due to the forced linear relationship in a non-linear space. An ordinal-trait aware SNP-phenotype regression may better model the skew in ordinal data commonly observed for traits ascertained through biobank surveys.^44^ Future work is necessary to understand the benefit of explicitly modeling ordinal data in genotype-phenotype associations of suicidal thoughts and behaviors. For example, to better understand how GWS loci increase or decrease risk for suicidality in a potentially non-linear fashion across suicidality categories.

To our knowledge, the enrichment of SNP-annotations related to conserved genomic regions is the first of its kind for traits along the suicidality spectrum but was consistent across approaches. The magnitude of enrichment also was consistent with those reported for major depressive disorder^40^ and across psychiatric disorders more broadly.^45^ In the context of loci identified in GWAS of major depression, genes found in conserved regions of the genome were part of networks relevant for organismal development and function across the lifetime such as synaptic function and brain development.^46, 47^ These enrichments in major depression GWAS suggest a rich interaction between genetic factors and the environment that has been empirically demonstrated for suicidality and select environments related to stress,^16^ substance use,^15^ and depression.^48^

Several tissue transcriptomic profiles from brain regions were nominally enriched in the GWAS of suicidality but the S-factor GWAS was underpowered to detect similar enrichments. We further tested these enrichments using a TWAS approach in the cerebellar hemisphere, cerebellum, hippocampus, and hypothalamus. There were extremely high correlations between the suicidality and S-factor regardless of tissue; however, these relationships were estimated using only *cis*-elements only.^38^ We therefore cannot rule out the contribution of *trans*-regulatory elements to gene expression differences between suicidality and the S-factor.

Though no gene reached genome-wide significance, the two most significantly associated genes harbor interesting functional relevance worth discussing. *RBM26* was associated with schizophrenia in a recent study but only at a level of suggestive significance (*P*=3.41×41^-7^).^49^ Within the first decade of diagnosis, people who suffer from schizophrenia are at the highest risk for suicidality, with a total suicide rate of 10%.^50^ Though there are several factors contributing to decreased life expectancy in schizophrenics, suicide is the largest one. *COLQ* is associated with cardiovascular traits such as resting heart rate (*P*=1.59×59^-12^).^51^ An increased baseline resting HR of 10 beats per minute increased the suicide rate by 19% in one study.^52^ Though accounting for many essential covariates such as smoking status, sex, body mass index, stress, depressed mood, and use of psychotropic medications, this study, to our knowledge, failed to account for population stratification and socioeconomic status. Though not directly related, *RBM26* and *COLQ* may have pleiotropic links to suicidality. Further research is required to untangle the cause-effect relationships between these potential risk factors and the severity of suicidal thoughts and behaviors.

We performed a cross-ancestry PGS analysis with respect suicidality and the S-factor which showed (i) limited portability to the AMR population with respect to suicidality measures, (ii) strong portability to the CSA population with respect to the S-factor indicator TLNWL, (iii) limited association between suicidality or S-factor PGS with other mental health outcomes associated with death by suicide, and (iv) lack of association between PGS and height, an unrelated trait. These findings are in line with the limited translation of EUR-derived PGS in existing cross-ancestry studies of transdiagnostic mental health characteristics.^53^ Of note from our study is the relatively large variance explained by the PGS in some of the diverse ancestries tested. This may partially be attributed to methodological benefits of a Bayesian approach but may also suggest consistent genetic architectures of suicidality across populations. Large studies are ongoing to learn about the genetic components of suicidal thoughts and behaviors in diverse ancestries and will permit deeper investigation of the S-factor and suicidality.

We demonstrated that the genetic and transcriptomic signatures of suicidality and the S-factor strongly overlap but our study has some limitations to consider. First, this body of work relies on large studies of European ancestry individuals clustered into this grouping using genetic principal components. Our results support limited translation of these results to diverse populations. It is well documented that these communities experience (i) social and cultural stigma surrounding suicidal thoughts, behaviors, and associated death^54, 55^ and (ii) vastly different face-to-face interactions with the healthcare system.^56^ For these reasons, our findings may not translate across diverse communities that disproportionately experience these thoughts and behaviors. Dedicated community outreach, sample recruitment, and educational programs are necessary to perform robust studies of suicidal thoughts and behaviors in other contexts, especially as they relate to community stressors that may interact with underlying genetic factors. Second, the UK Biobank is limited by the potential for recruitment, survivor, and recall bias as this cohort is generally older, wealthier, and better educated than a general community sampling. This cohort may therefore be depleted for the more extreme ends of the suicidality spectrum and better reflect milder suicidality ratings than those from a more representative sampling. Finally, death by suicide and the preceding thoughts and behaviors routinely co-occur with psychiatric diagnoses. The GWAS used to construct the S-factor did not take into consideration the effects of co-morbid depression, anxiety, chronic pain, or other necessary experiences or diagnoses. There is evidence that genetic findings related to suicidality are independent of psychiatric diagnoses^1, 6, 7, 11^ but it remains unclear how best to account for these variables to make discoveries with as much specificity for suicidal thoughts and behaviors as possible. Finally, we used dichotomous measures of suicidality as indicator traits to construct the S-factor. The construction of these items (e.g., the suicidal and non-suicidal self-injury in the ASH phenotype) may underestimate heritability and its contribution to the S-factor structure.

Despite these limitations, this study empirically investigated the differences and similarities between definitions of suicidality. Across all analyses presented, the phenotypically-aggregated suicidality item was more statistically powerful and informative than the S-factor for downstream *in silico* characterization of the biology underlying this complex trait. In conclusion, our study informs one path forward for the analysis of participant responses related to thoughts and behaviors associated with death by suicide. By aggregating multiple informative items into a suicidality phenotype, studies are likely to generate more information about suicide biology compared to individual binary items or a genetically-defined S-factor.

## Data Availability

This research was conducted using the UK Biobank Resource (application reference no. 58146). UKB data can be accessed by bona fide researchers through the UKB data access portal.
Suicidality GWAS data can be downloaded here: https://researchdata.gla.ac.uk/930/
S-factor GWAS data will be shared via secure zenodo link at the time of publication.
All other data produced in the present work are contained in the manuscript.

## ACKNOWLEDGEMENTS

This research was conducted using the UK Biobank Resource (application reference no. 58146). The authors thank the research participants and employees of the UK Biobank for making this work possible. This study was supported in part by the National Institutes of Health (R21DC018098 and R33DA047527) and One Mind.

## SUPPLEMENTARY MATERIAL

**Table S1**. SNP-based heritability estimates for the four indicator traits used to derive the S-factor.

**Table S2**. Genetic correlation estimates (s.e.) for the four indicator traits used to derive the S-factor.

**Table S3**. Structure of the S-factor using two separate models. Model 1 includes attempted suicide in the factor structure while model 2 removes it due to lack of significant SNP-based heritability estimate.

**Table S4**. Comparison of effect sizes for three prior suicidality SNPs discovered in Strawbridge, et al.^11^

**Table S5**. Summary of effect size distribution comparisons between suicidality and the S-factor. Table A compares effect size distribution metrics; B compares the projected number of SNPs recovered from increasing sample sizes of each trait; C compares the projected variance explained from increasing sample sizes of each trait.

**Table S6**. Partitioned heritability results for enrichment of genomic annotations. Yellow boxes denote significant enrichments.

**Table S7**. Genetic correlation estimates for 32 mental health traits related to suicidality and the S-factor.

**Table S8**. Tissue transcriptomic profile enrichment for suicidality and the S-factor using GTEx v8.

**Table S9**. TWAS results for suicidality and S-factor in four brain tissues (GTEx v8): cerebellar hemisphere, cerebellum, hippocampus, and hypothalamus.

**Table S10**. Results of cross-population polygenic scoring. Yellow highlight indicates a significant result in either the suicidality test or the S-factor test. Yellow highlight plus red text indicates a significant result in both tests.

## REFERENCES

1. Mullins N, Kang J, Campos AI, Coleman JRI, Edwards AC, Galfalvy H et al. Dissecting the Shared Genetic Architecture of Suicide Attempt, Psychiatric Disorders, and Known Risk Factors. Biol Psychiatry 2022; 91(3): 313–327.

2. F World Health Organization Suicide Facto Sheet. https://www.who.int/news-room/fact-sheets/detail/suicide. Accessed July 14, 2022.

3. World Health Organization: Preventing Suicide: A Global Perspective. http://apps.who.int/iris/bitstream/handle/10665/131056/9789241564779_eng.pdf;jsessionid=. Accessed July 14, 2022.

4. Kimbrel NA, Ashley-Koch AE, Qin XJ, Lindquist JH, Garrett ME, Dennis MF et al. A genome-wide association study of suicide attempts in the million veterans program identifies evidence of pan-ancestry and ancestry-specific risk loci. Molecular Psychiatry 2022; 27(4): 2264–2272.

5. Erlangsen A, Appadurai V, Wang Y, Turecki G, Mors O, Werge T et al. Genetics of suicide attempts in individuals with and without mental disorders: a population-based genome-wide association study. Molecular Psychiatry 2020; 25(10): 2410–2421.

6. Docherty AR, Mullins N, Ashley-Koch AE, Qin XJ, Coleman J, Shabalin AA et al. Genome-wide association study meta-analysis of suicide attempt in 43,871 cases identifies twelve genome-wide significant loci. medRxiv 2022: 2022.2007.2003.22277199.

7. Mullins N, Bigdeli TB, Børglum AD, Coleman JRI, Demontis D, Mehta D et al. GWAS of Suicide Attempt in Psychiatric Disorders and Association With Major Depression Polygenic Risk Scores. Am J Psychiatry 2019; 176(8): 651–660.

8. Stein MB, Levey DF, Cheng Z, Wendt FR, Harrington K, Pathak GA et al. Genome-wide association analyses of post-traumatic stress disorder and its symptom subdomains in the Million Veteran Program. Nature Genetics 2021; 53(2): 174–184.

9. Wendt FR, Pathak GA, Deak JD, De Angelis F, Koller D, Cabrera-Mendoza B et al. Using phenotype risk scores to enhance gene discovery for generalized anxiety disorder and posttraumatic stress disorder. Mol Psychiatry 2022; 27(4): 2206–2215.

10. Levey DF, Stein MB, Wendt FR, Pathak GA, Zhou H, Aslan M et al. Bi-ancestral depression GWAS in the Million Veteran Program and meta-analysis in >1.2 million individuals highlight new therapeutic directions. Nature Neuroscience 2021; 24(7): 954–963.

11. Strawbridge RJ, Ward J, Ferguson A, Graham N, Shaw RJ, Cullen B et al. Identification of novel genome-wide associations for suicidality in UK Biobank, genetic correlation with psychiatric disorders and polygenic association with completed suicide. EBioMedicine 2019; 41: 517–525.

12. Grotzinger AD, Mallard TT, Akingbuwa WA, Ip HF, Adams MJ, Lewis CM et al. Genetic architecture of 11 major psychiatric disorders at biobehavioral, functional genomic and molecular genetic levels of analysis. Nat Genet 2022; 54(5): 548–559.

13. Grotzinger AD, Rhemtulla M, de Vlaming R, Ritchie SJ, Mallard TT, Hill WD et al. Genomic structural equation modelling provides insights into the multivariate genetic architecture of complex traits. Nat Hum Behav 2019; 3(5): 513–525.

14. Mallard TT, Savage JE, Johnson EC, Huang Y, Edwards AC, Hottenga JJ et al. Item-Level Genome-Wide Association Study of the Alcohol Use Disorders Identification Test in Three Population-Based Cohorts. Am J Psychiatry 2022; 179(1): 58–70.

15. Polimanti R, Levey DF, Pathak GA, Wendt FR, Nunez YZ, Ursano RJ et al. Multi-environment gene interactions linked to the interplay between polysubstance dependence and suicidality. Transl Psychiatry 2021; 11(1): 34.

16. Wendt FR, Pathak GA, Levey DF, Nuñez YZ, Overstreet C, Tyrrell C et al. Sex-stratified gene-by-environment genome-wide interaction study of trauma, posttraumatic-stress, and suicidality. Neurobiol Stress 2021; 14: 100309.

17. Finucane HK, Bulik-Sullivan B, Gusev A, Trynka G, Reshef Y, Loh PR et al. Partitioning heritability by functional annotation using genome-wide association summary statistics. Nat Genet 2015; 47(11): 1228–1235.

18. Gazal S, Marquez-Luna C, Finucane HK, Price AL. Reconciling S-LDSC and LDAK functional enrichment estimates. Nat Genet 2019; 51(8): 1202–1204.

19. Koller D, Wendt FR, Pathak GA, De Lillo A, De Angelis F, Cabrera-Mendoza B et al. The impact of evolutionary processes in shaping the genetics of complex traits in East Asia and Europe: a specific contribution from Denisovan and Neanderthal introgression. bioRxiv 2021: 2021.2008.2012.456138.

20. Wendt FR, Pathak GA, Overstreet C, Tylee DS, Gelernter J, Atkinson EG et al. Characterizing the effect of background selection on the polygenicity of brain-related traits. Genomics 2021; 113(1 Pt 1): 111–119.

21. Demontis D, Walters RK, Martin J, Mattheisen M, Als TD, Agerbo E et al. Discovery of the first genome-wide significant risk loci for attention deficit/hyperactivity disorder. Nat Genet 2019; 51(1): 63–75.

22. Watson HJ, Yilmaz Z, Thornton LM, Hübel C, Coleman JRI, Gaspar HA et al. Genome-wide association study identifies eight risk loci and implicates metabo-psychiatric origins for anorexia nervosa. Nat Genet 2019; 51(8): 1207–1214.

23. Revealing the complex genetic architecture of obsessive-compulsive disorder using meta-analysis. Mol Psychiatry 2018; 23(5): 1181–1188.

24. Trubetskoy V, Pardiñas AF, Qi T, Panagiotaropoulou G, Awasthi S, Bigdeli TB et al. Mapping genomic loci implicates genes and synaptic biology in schizophrenia. Nature 2022; 604(7906): 502–508.

25. Yu D, Sul JH, Tsetsos F, Nawaz MS, Huang AY, Zelaya I et al. Interrogating the Genetic Determinants of Tourette’s Syndrome and Other Tic Disorders Through Genome-Wide Association Studies. Am J Psychiatry 2019; 176(3): 217–227.

26. Zhou H, Sealock JM, Sanchez-Roige S, Clarke TK, Levey DF, Cheng Z et al. Genome-wide meta-analysis of problematic alcohol use in 435,563 individuals yields insights into biology and relationships with other traits. Nat Neurosci 2020; 23(7): 809–818.

27. Levey DF, Gelernter J, Polimanti R, Zhou H, Cheng Z, Aslan M et al. Reproducible Genetic Risk Loci for Anxiety: Results From ∼200,000 Participants in the Million Veteran Program. Am J Psychiatry 2020; 177(3): 223–232.

28. de Moor MH, Costa PT, Terracciano A, Krueger RF, de Geus EJ, Toshiko T et al. Meta-analysis of genome-wide association studies for personality. Mol Psychiatry 2012; 17(3): 337–349.

29. Okbay A, Baselmans BM, De Neve JE, Turley P, Nivard MG, Fontana MA et al. Genetic variants associated with subjective well-being, depressive symptoms, and neuroticism identified through genome-wide analyses. Nat Genet 2016; 48(6): 624–633.

30. Karlsson Linnér R, Biroli P, Kong E, Meddens SFW, Wedow R, Fontana MA et al. Genome-wide association analyses of risk tolerance and risky behaviors in over 1 million individuals identify hundreds of loci and shared genetic influences. Nat Genet 2019; 51(2): 245–257.

31. Lee JJ, Wedow R, Okbay A, Kong E, Maghzian O, Zacher M et al. Gene discovery and polygenic prediction from a genome-wide association study of educational attainment in 1.1 million individuals. Nat Genet 2018; 50(8): 1112–1121.

32. Zhang Y, Qi G, Park J-H, Chatterjee N. Estimation of complex effect-size distributions using summary-level statistics from genome-wide association studies across 32 complex traits. Nature Genetics 2018; 50(9): 1318–1326.

33. Johnson EC, Kapoor M, Hatoum AS, Zhou H, Polimanti R, Wendt FR et al. Investigation of convergent and divergent genetic influences underlying schizophrenia and alcohol use disorder. Psychol Med 2021: 1–9.

34. Yengo L, Vedantam S, Marouli E, Sidorenko J, Bartell E, Sakaue S et al. A Saturated Map of Common Genetic Variants Associated with Human Height from 5.4 Million Individuals of Diverse Ancestries. bioRxiv 2022: 2022.2001.2007.475305.

35. de Leeuw CA, Mooij JM, Heskes T, Posthuma D. MAGMA: Generalized Gene-Set Analysis of GWAS Data. PLOS Computational Biology 2015; 11(4): e1004219.

36. Watanabe K, Taskesen E, van Bochoven A, Posthuma D. Functional mapping and annotation of genetic associations with FUMA. Nature Communications 2017; 8(1): 1826.

37. Gusev A, Ko A, Shi H, Bhatia G, Chung W, Penninx BW et al. Integrative approaches for large-scale transcriptome-wide association studies. Nat Genet 2016; 48(3): 245–252.

38. Mancuso N, Shi H, Goddard P, Kichaev G, Gusev A, Pasaniuc B. Integrating Gene Expression with Summary Association Statistics to Identify Genes Associated with 30 Complex Traits. Am J Hum Genet 2017; 100(3): 473–487.

39. Ge T, Chen CY, Ni Y, Feng YA, Smoller JW. Polygenic prediction via Bayesian regression and continuous shrinkage priors. Nat Commun 2019; 10(1): 1776.

40. Howard DM, Adams MJ, Clarke TK, Hafferty JD, Gibson J, Shirali M et al. Genome-wide meta-analysis of depression identifies 102 independent variants and highlights the importance of the prefrontal brain regions. Nat Neurosci 2019; 22(3): 343–352.

41. Zhang Y, Qi G, Park JH, Chatterjee N. Estimation of complex effect-size distributions using summary-level statistics from genome-wide association studies across 32 complex traits. Nat Genet 2018; 50(9): 1318–1326.

42. Davis KAS, Coleman JRI, Adams M, Allen N, Breen G, Cullen B et al. Mental health in UK Biobank - development, implementation and results from an online questionnaire completed by 157 366 participants: a reanalysis. BJPsych Open 2020; 6(2): e18.

43. Trubetskoy V, Pardiñas AF, Qi T, Panagiotaropoulou G, Awasthi S, Bigdeli TB et al. Mapping genomic loci implicates genes and synaptic biology in schizophrenia. Nature 2022; 604(7906): 502–508.

44. Bi W, Zhou W, Dey R, Mukherjee B, Sampson JN, Lee S. Efficient mixed model approach for large-scale genome-wide association studies of ordinal categorical phenotypes. Am J Hum Genet 2021; 108(5): 825–839.

45. Romero C, Werme J, Jansen PR, Gelernter J, Stein MB, Levey D et al. Exploring the genetic overlap between 12 psychiatric disorders. medRxiv 2022: 2022.2004.2012.22273763.

46. Sall S, Thompson W, Santos A, Dwyer DS. Analysis of Major Depression Risk Genes Reveals Evolutionary Conservation, Shared Phenotypes, and Extensive Genetic Interactions. Frontiers in Psychiatry 2021; 12.

47. Wray NR, Ripke S, Mattheisen M, Trzaskowski M, Byrne EM, Abdellaoui A et al. Genome-wide association analyses identify 44 risk variants and refine the genetic architecture of major depression. Nature Genetics 2018; 50(5): 668–681.

48. Coleman JRI, Peyrot WJ, Purves KL, Davis KAS, Rayner C, Choi SW et al. Genome-wide gene-environment analyses of major depressive disorder and reported lifetime traumatic experiences in UK Biobank. Mol Psychiatry 2020; 25(7): 1430–1446.

49. Pardiñas AF, Holmans P, Pocklington AJ, Escott-Price V, Ripke S, Carrera N et al. Common schizophrenia alleles are enriched in mutation-intolerant genes and in regions under strong background selection. Nat Genet 2018; 50(3): 381–389.

50. Sher L, Kahn RS. Suicide in Schizophrenia: An Educational Overview. Medicina (Kaunas) 2019; 55(7).

51. Zhu Z, Wang X, Li X, Lin Y, Shen S, Liu CL et al. Genetic overlap of chronic obstructive pulmonary disease and cardiovascular disease-related traits: a large-scale genome-wide cross-trait analysis. Respir Res 2019; 20(1): 64.

52. Lemogne C, Thomas F, Consoli SM, Pannier B, Jégo B, Danchin N. Heart rate and completed suicide: evidence from the IPC cohort study. Psychosom Med 2011; 73(9): 731–736.

53. Docherty AR, Moscati A, Dick D, Savage JE, Salvatore JE, Cooke M et al. Polygenic prediction of the phenome, across ancestry, in emerging adulthood. Psychol Med 2018; 48(11): 1814–1823.

54. Ammerman BA, Fahlgren MK, Sorgi KM, McCloskey MS. Differences in Suicidal Thoughts and Behaviors Among Three Racial Groups. Crisis 2020; 41(3): 172–178.

55. Colucci E, Lester D. A cross-cultural study of attitudes toward suicide among young people in India, Italy and Australia. Int J Soc Psychiatry 2020; 66(7): 700–706.

56. Karen J. Coleman, Ph.D.,, Christine Stewart, Ph.D.,, Beth E. Waitzfelder, Ph.D.,, John E. Zeber, Ph.D.,, Leo S. Morales, M.D., Ph.D.,, Ameena T. Ahmed, M.D., M.P.H., et al. Racial-Ethnic Differences in Psychiatric Diagnoses and Treatment Across 11 Health Care Systems in the Mental Health Research Network. Psychiatric Services 2016; 67(7): 749–757.

